# How COVID-19 continues to affect contraception in Scotland: a retrospective analysis of Scottish prescribing data between 2016 and 2023

**DOI:** 10.1101/2023.09.14.23295542

**Authors:** Elliot Johnson-Hall

## Abstract

**Background:** In Scotland, the effects of the COVID-19 pandemic on women’s access to contraception are unknown. Globally, COVID-19 restrictions have led to a shift to telehealth service delivery alongside a reduction in contraceptive provision. Research into whether the effects of COVID-19 on contraception have abated after restrictions have been lifted is lacking.

**Methods:** This is a retrospective longitudinal study of prescribing data from the Scottish Health and Social Care Open Data repository (https://www.opendata.nhs.scot) between January 2016 and January 2023. Contraceptives were extracted and categorised using truncated British National Formulary codes and analysed using R. Contraceptive provision was compared across four periods: pre-COVID-19 (01/01/2016–23/03/2020), lockdown (24/03/2020–29/05/2020 & 05/01/2021–26/04/2021), restrictions (30/05/2020–04/01/2021 & 27/04/2021–30/04/2022), and post-COVID-19 (01/05/2022–01/01/2023).

**Results:** During lockdowns, contraceptive prescribing in Scotland decreased by 82.90% of pre-COVID-19 levels. This trend was more severe for long-acting reversible contraception which fell to 11.80% of pre-COVID-19 prescriptions. After COVID-19, the level of contraceptive prescribing has risen to 108.23% of its pre-pandemic level. Large increases in subcutaneous medroxyprogesterone acetate (499.05%), progestogen-only pills (125.07%), the patch (165.09%), levonorgestrel-IUS (112.54%), and ulipristal acetate emergency contraception prescribing (357.97%). Conversely, combined oral contraceptive pills (75.04%), Cu-IUD (83.63%), the implant (81.10%), and levonorgestrel emergency contraception (67.42%) prescribing has decreased.

**Conclusions:** COVID-19 vastly decreased contraceptive prescribing during lockdowns in Scotland. Post-COVID-19, changes in contraceptive prescribing within Scottish general practices are reported, with implications for health policy and service delivery planning.

**Availability of Data & Code:** All code and data used are fully available from Zenodo (doi:10.5281/zenodo.8310085)

The raw dataset used is also publicly available from the Scottish Health and Social Care Open Data repository (opendata.nhs.scot).

## Introduction

COVID-19 lockdowns decreased contraception dispensed in England [1]. Furthermore, women in the UK found accessing contraception more difficult due to COVID-19 [2]. This is not a trend unique to the UK, but a global phenomenon [3–5].

Approximately 75% of Scottish women access contraception through their general practice (GP) [6]. Restrictions due to COVID-19 changed day-to-day life in many ways, including the format of GP consultations. To mitigate the risk of COVID-19 infection, GPs pivoted to remote consultations such as telephone triage and video-based telehealth appointments wherever possible [7], the basis for which was established in Scotland [8].

The Scottish Government expected that GPs would be unable to deliver some services due to increased demand [9]. Moreover, the Faculty of Sexual & Reproductive Healthcare (FSRH) guidance issued during the pandemic did not classify the provision of oral contraception or new long-acting reversible contraception (LARC) fittings as essential services needing face-to-face appointments [10].

Despite this, no study has yet examined the effects of COVID-19 on contraceptive provision in Scotland. Additionally, much of the literature published has not assessed the changes on contraceptive prescribing now the acute phase of the pandemic has passed. Although, a forthcoming systematic review may illuminate these issues [11].

This study will examine the effects of COVID-19 lockdowns and restrictions on the provision of contraception in Scottish general practices and how contraceptive provision has changed in post-COVID-19 Scotland.

## Materials and methods

This longitudinal retrospective study examines the period between January 2016 and January 2023, utilising data from the Scottish Health and Social Care Open Data repository (https://www.opendata.nhs.scot). This aggregated, anonymous dataset is 100% complete for all prescriptions dispensed outside hospitals throughout Scotland. This dataset is published monthly; thus, observing changes in day-to-day prescribing is impossible.

The majority of prescriptions in this dataset are prescribed in general practices and dispensed by community pharmacies. Items dispensed in hospitals, prisons, schools, and private prescriptions, as well as prescriptions not presented for dispensing and those dispensed but not submitted for payment are excluded. These exclusions are unlikely to be a large enough proportion of these data to impact the observed trends [12].

R v4.3.0 [13] was used to create a script (available from Zenodo: doi.org/10.5281/zenodo.8310084) to access data from the NHS Scotland Open Data API. Initially, the complete dataset is filtered using a SQL query to extract and categorise contraceptive medicines by returning results with truncated British National Formulary (BNF) [14] item codes beginning with 07030* or 21040* (Table 1).

**Table 1.**
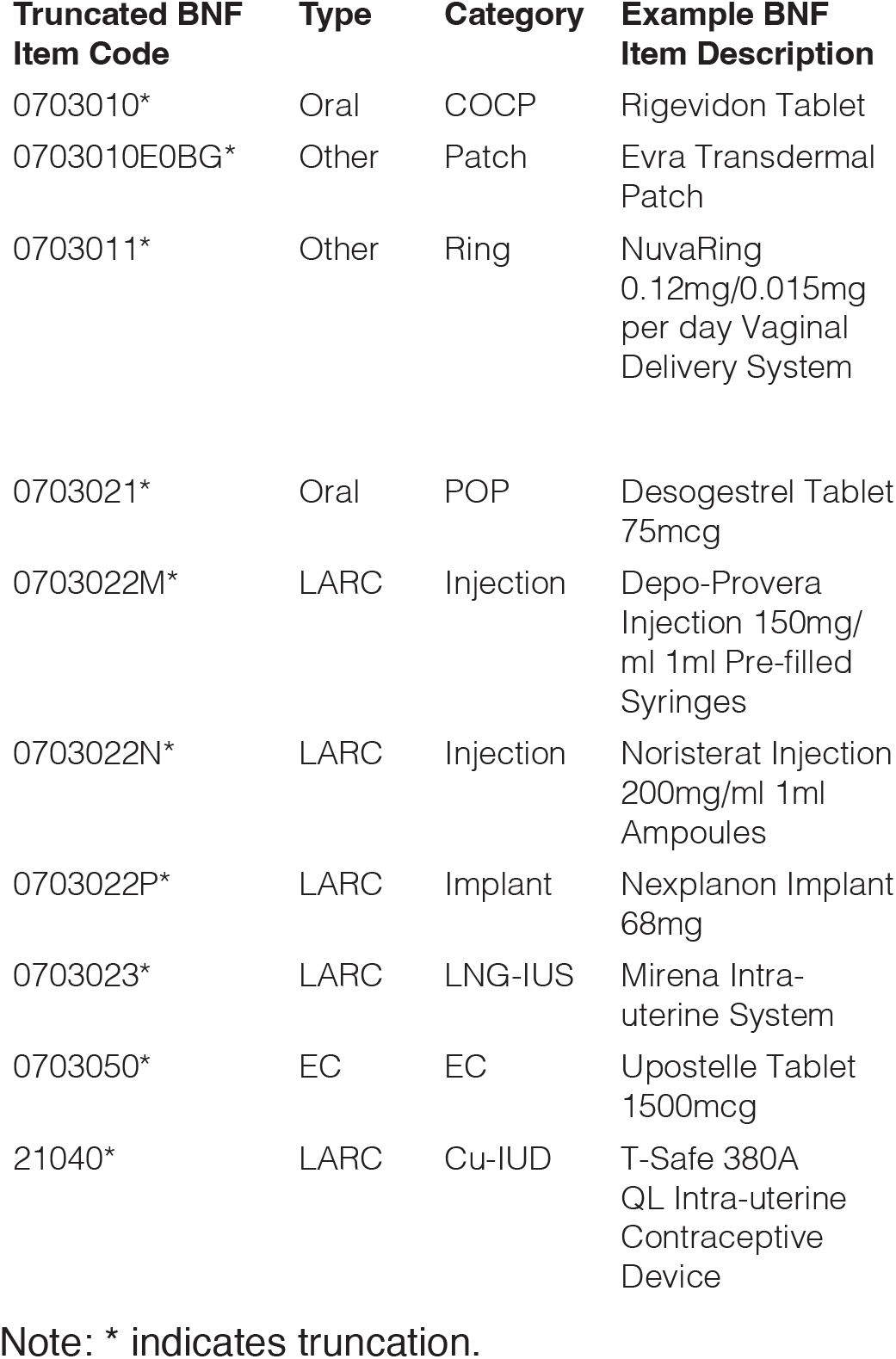
Truncated BNF item codes used during data extraction. Type, categories, and example medicines in for each code.

Spermicidal jelly (e.g. Nonoxinol) was removed from this analysis as it is not a contraceptive in its own right as it needs to be employed with a barrier method [14]. As only 0.012% (64,647 / 532,181,153) of the total items dispensed in the duration of this study, the exclusion of this method of contraception did not affect the conclusions drawn.

Due to inherent differences in prescribing frequencies between contraceptive methods, a standardised metric—’months of contraceptive coverage’ (MCC)— was calculated:

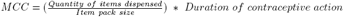

For example, a Cu-IUD with a five-year lifespan provides 60 months of contraceptive coverage (1/1 * 60). In contrast, a six-month prescription of a short-acting oral contraceptive offers six months of contraceptive coverage (126/21 * 1) despite 126 items being dispensed.

To compare prescribing trends between periods, mean months of contraceptive coverage dispensed (MMCC) was calculated by dividing the months of contraceptive coverage by the number of months in each period. However, emergency contraception (EC) was compared by the mean number of items dispensed per month.

Four different periods were defined. Firstly, pre-COVID-19 (01/01/2016–23/03/2020). Lockdown (24/03/2020–29/05/2020 and 05/01/2021– 26/04/2021) when Scotland entered the highest level of restrictions on daily activities [15]. Thirdly, restrictions (30/05/2020–04/01/2021 and 27/04/2021–30/04/2022) when - due to its geographic diversity - Scotland adopted a phased approach to restrictions based on local epidemiological data [16]. Thus, during this period, Scotland experienced a variety of restrictions on both local, regional, and national levels. Finally, post-COVID-19 (01/05/2022– 01/01/2023), when all restrictions in Scotland were lifted [15].

## Results

### Overview

During lockdown, total contraceptive prescribing in Scotland fell to 17.10% (98,642 MMCC) of pre-COVID-19 levels (576,986 MMCC). Between lockdowns, prescribing of all forms of contraception recovered to 76.96% (444,055 MMCC) of pre-COVID-19 levels. Intriguingly, the post-COVID-19 period has seen an increase in overall contraceptive prescribing compared with pre-COVID-19 (108.23% / 624,474 MMCC).

The large decrease in contraceptive provision during periods of COVID-19 lockdowns was seen across all forms of contraception: LARC fell to 11.80% (19,424 MMCC), oral to 17.64% (54,852 MMCC), the ring and patch to 24.01% (24,366 MMCC). EC also dropped to 13.90% of pre-COVID-19 supply (1,118 dispensed).

Restrictions allowed the resumption of contraceptive dispensing, albeit at a lower rate than pre-COVID-19: LARC: 64.40% (105,964 MMCC), oral 74.91% (232,923 MMCC), EC 84.60% (6,804 dispensed). However, for the ring and patch there was an overall increase (103.62% / 105,168 MMCC) in dispensing during periods of COVID-19 restrictions compared with pre-COVID-19.

Overall, oral contraceptives remained the majority of prescribed contraceptives throughout the duration of this study. 52.67% (903,036 MMCC) of the total dispensed (1,744,157 MMCC). Behind this, LARC (24.48% / 451,312 MMCC), and finally the contraceptive patch and ring (22.85% / 389,909 MMCC). However, the proportions and types of contraceptives prescribed in each of the four periods of this study varied (Figure 1).

**Figure 1.**
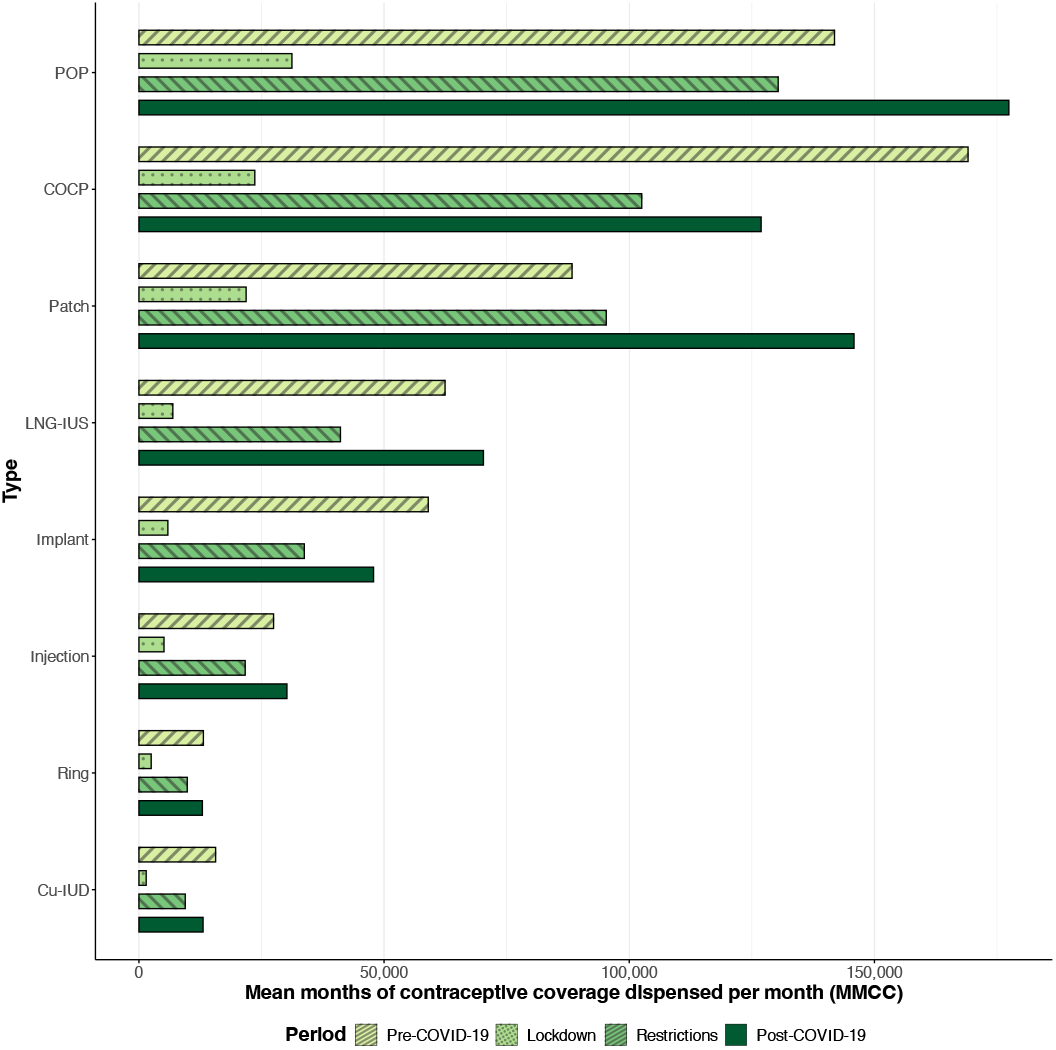
COVID-19 restrictions changed the proportions of categories of contraception dispensed in Scotland. Prior to COVID-19, the combined oral contraceptive pill (COCP), was the most dispensed form of contraception. However, during the COVID-19 pandemic, this changed to the progestogen-only pill (POP), with dispensing of the contraceptive patch also increasing. Dispensing rates of long-acting reversible contraception (LARC), including the LNG-IUS, Cu-IUD, injection, and implant decreased during COVID-19, whilst the contraceptive ring remained relatively stable. Post-COVID-19 data indicates these changes in contraceptive prescribing persist in Scottish general practices.

### Oral contraception

Before the COVID-19 pandemic, combined oral contraceptives were more frequently prescribed (mean percentage of total MMCC of oral contraception dispensed: 54.38%) than progestogen-only contraceptives (45.62%). However, during the period from April 2020 to April 2022, this trend reversed (COCP: 44.03%, POP: 55.97%).

Intriguingly, the trend of decreasing COCP and increasing POP in Scotland has continued (COCP: 41.70%, POP: 58.30%), even after COVID-19-associated restrictions were lifted (Figure 2).

**Figure 2.**
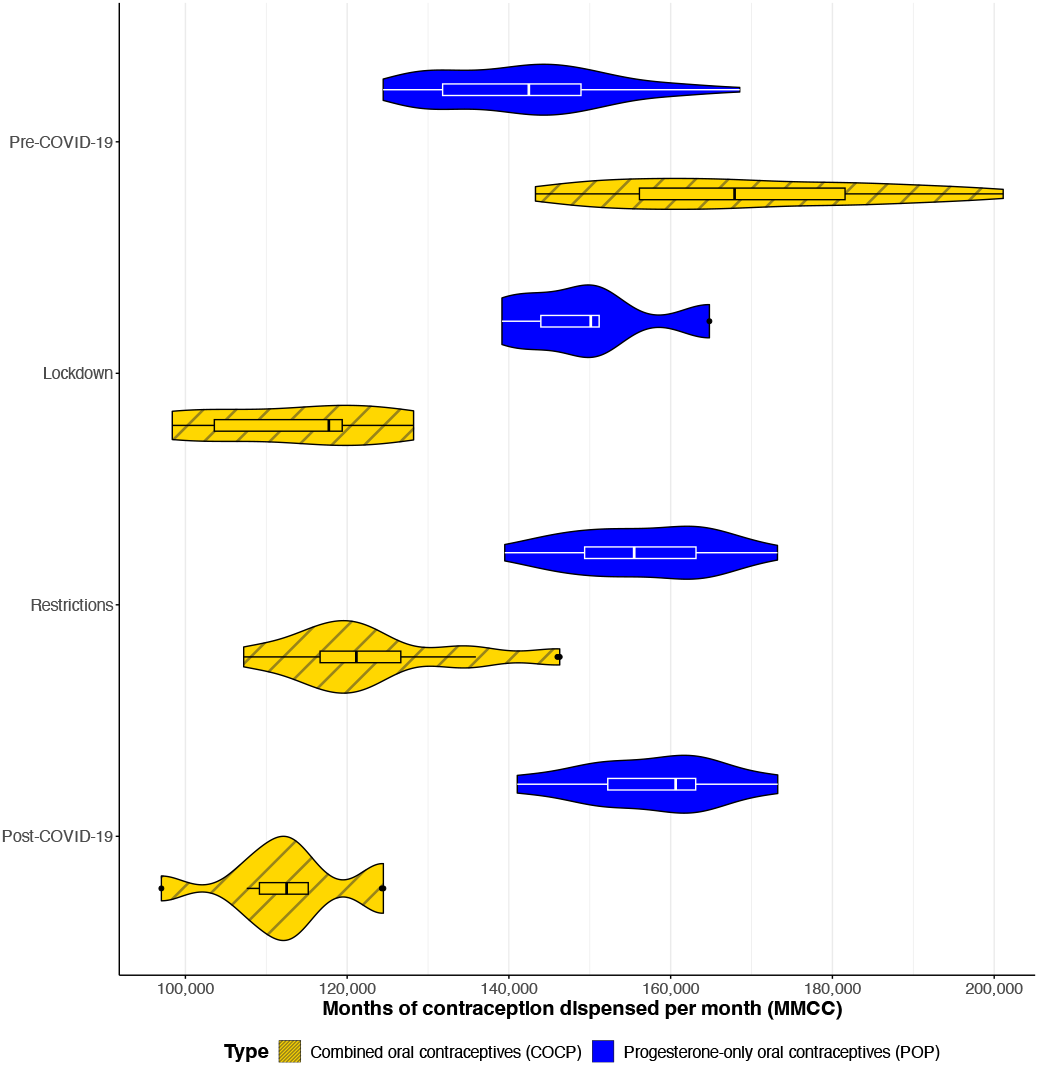
Changes in oral contraceptive prescribing. Prior to COVID-19, COCPs were more prescribed than POPs. However, there was a rough balance between the two methods. During lockdown, POPs were vastly more prescribed, and COCPs decreased. This trend continued during periods of restrictions, and after COVID-19, this gap has widened.

### Long-acting reversible contraception

Long-acting reversible contraception (LARC) requires administration by a healthcare professional [17,22,23] – with the exception of the Sayana-Press (depot medroxyprogesterone acetate) subcutaneous injection [24]. Thus, due to the restrictions on face-to-face appointments, LARC administration severely decreased throughout the COVID-19 pandemic (Figure 3).

**Figure 3.**
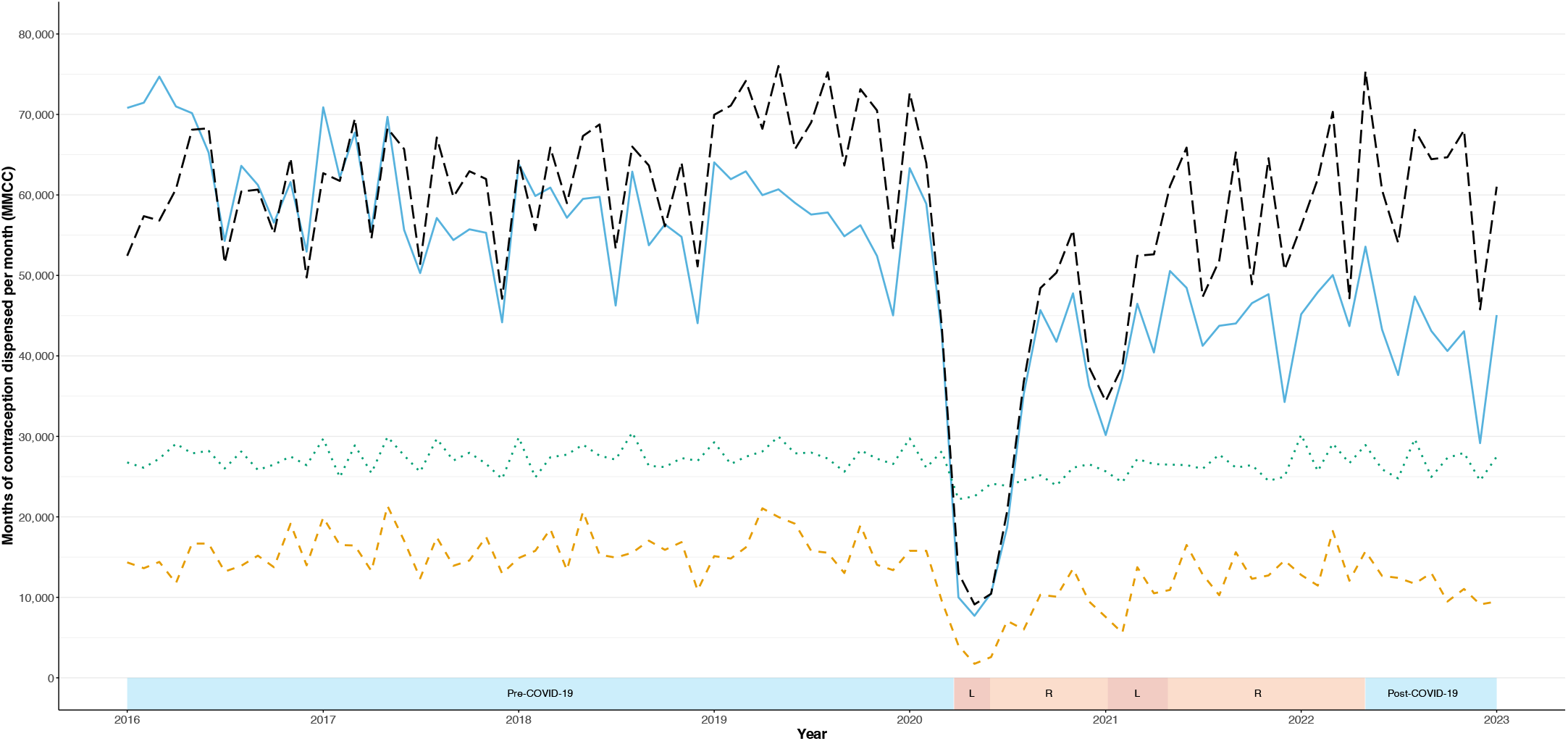
LARC provision by time period. Long-acting reversible contraceptive (LARC) prescribing fell enormously during the first lockdown (red-shaded periods marked L). During periods of restrictions (orange-shaded periods, R) still hampered access to LARC. Post-COVID-19, LARC prescribing is slightly reduced compared with pre-COVID-19.

### Cu-IUD & LNG-IUS

During lockdowns, levonorgestrel intrauterine system (LNG-IUS) prescribing sank to 11.07% (6,912 MMCC) of pre-COVID-19 levels, while copper intrauterine device (Cu-IUD) prescribing decreased to 9.48% (1,482 MMCC). Subsequently, during periods of COVID-19-associated restrictions, LNG-IUS prescribing increased to 65.84% (41,102 MMCC) and Cu-IUD to 60.45% (9,455 MMCC). Post-COVID-19, Cu-IUD prescribing recovered to 83.63% (13,080 MMCC) of pre-COVID-19 levels. Conversely, LNG-IUS prescribing post-COVID-19 has exceeded that before the pandemic, reaching 112.54% (70,260 MMCC) of pre-COVID-19 levels.

### Implant

Provision of the progestogen-only contraceptive implant fell to 10.02% (5913 MMCC) during lockdown. Dispensing rebounded to 57.17% (33,730 MMCC) during periods of restrictions. Post-COVID-19 implant prescribing remains at 81.10% (47,853 MMCC) of pre-COVID-19 levels (59,004 MMCC).

### Injection

The decrease in injection dispensing throughout COVID-19 is less severe than other forms of LARC during periods of lockdown (18.63% / 5,116 MMCC) and restrictions (78.92% / 21,676 MMCC) compared with pre-COVID-19 levels (27,466 MMCC). However, there was a change in route of administration, with intramuscular (IM) Depo-Provera decreasing to 15.12% (3,927 MMCC) of pre-pandemic levels (25,979 MMCC) during periods of lockdown and subcutaneous (SC) Sayana Press decreasing to 80.06% (1189 / 1485 MMCC).

During periods of restrictions, the difference between SC and IM injections becomes obvious. IM injection during restrictions fell to 63.83% (16,583 MMCC) of pre-COVID-19 levels, whereas SC injections rose to 343.04% (5,093 MMCC). Post-COVID-19 this trend has increased, with IM prescribing at 87.68% (22,779 MMCC) and SC at 499.05% (7,409 MMCC) of pre-COVID-19 levels. Whilst IM injections are still more prescribed, SC injections are rapidly increasing in popularity.

### Patch

The patch is unique among the categories of contraception assessed here as it increased in popularity (107.89% / 95,311 MMCC) compared with pre-COVID-19 levels (88,340 MMCC) during the period of restrictions and has increased again in the post-COVID-19 period (165.09% / 145,840 MMCC). However, patch prescribing did reduce in lockdown to 24.75% (21,865 MMCC) of pre-COVID-19 levels.

### Emergency contraception

Before COVID-19, the mean number of emergency contraception (EC) items dispensed monthly was 8,043. Due to social restrictions present in Scotland, emergency contraception dispensing was expected to decrease. This was the case during both periods of restrictions (84.60% / 6,804 mean number of items dispensed per month) and lockdowns (13.90% / 1,118 items).

Another interesting observation is the change in the most dispensed form of EC from levonorgestrel to ulipristal acetate (54.61% / 5,320) post-COVID-19. Prior to COVID-19, 18.48% (1,486 items) of EC dispensed was ulipristal acetate, with the remainder being levonorgestrel. Ulipristal acetate dispensing increased during periods of lockdown to 39.98% (447) and during restrictions to 43.34% (2,949).

## Discussion

COVID-19 restrictions prevented the regular BMI and blood pressure monitoring required for safe COCP prescribing [17]. This likely led to the decrease seen in COCP prescribing and the growth in dispensing of POPs instead, which do not require the same patient monitoring [17–19].

Additionally, some of the increase in POP prescribing is likely due to the use of desogestrel as a bridging contraceptive due to extended LARC use during COVID-19 restrictions [20].

This may also reflect increased access to short-term supplies of POPs without a prescription within community pharmacies in Scotland [21]. Women who – due to COVID-19 restrictions - were unable to initiate prescribed contraception may have obtained contraception through community pharmacists, increasing POP dispensing. Continuing prescribing POP initiated through this route would at least partially explain this trend. However, the mechanism behind this increase is a clear area for further research.

The changes seen in injections can only be due to the fact Sayana Press subcutaneous injections are licenced for self-administration [24], as both contain the same active ingredient (medroxyprogesterone acetate) and last for at least 12 weeks (Depo-Provera) or 13 weeks (Sayana Press) [24,26] – a negligible difference.

The increase in EC dispensing post-COVID-19 (121.11% / 9,741 items) versus pre-COVID-19 levels potentially points to a lack of effective regular contraception due to disrupted access during COVID-19. However, it is impossible to state this with certainty, as other factors may underlie this trend. Emergency contraception is available free of charge without a prescription from most Scottish community pharmacies [27].

There are a variety of factors which may have led to this change. Patients could be presenting later after unprotected sexual intercourse which would make ulipristal acetate dispensing more likely [28]. However, this warrants further investigation.

## Conclusions

The clear decreases in contraceptive provision in Scotland during COVID-19 lockdowns present many possible outcomes ranging from unintended pregnancies to increases in terminations. The effects of these restrictions have altered trends in contraceptive prescribing within Scottish general practice.

Telemedicine-based contraceptive prescribing has been demonstrated to be effective and safe at a scale unthought-of before COVID-19. Within Scotland, this has the principal advantage of easier access to sexual and reproductive healthcare professionals, especially for those women living in island and rural communities. Additionally, this may increase the ease of access to Gaelic-speaking healthcare providers for Gaelic-speaking patients.

However, the potential lack of patient autonomy in choosing their preferred method of contraception, particularly LARC, during this period is understandable in the context of a global health emergency but troubling. Sexual healthcare services must learn from this period and work to ensure continuity of safe access to contraception can continue, despite external factors.

### Limitations

A key assumption is made that the patient population seeking contraception from general practitioners is constant throughout the period of this study, and consequently, variances in prescribing rates between pre-COVID-19 and post-COVID-19 are due to changes in contraceptive prescribing rather than population-level alterations.

It is also assumed that the drugs dispensed here are used purely for their licenced indication of contraception, not any alternative off-label uses. However, it is not possible to exclude medicines dispensed for non-contraceptive uses. Regardless, off-label use likely constitutes a minute proportion of this dataset.

Finally, whilst there is a period of over four years for the pre-COVID-19 data, only seven months of post-COVID-19 was available for this analysis.

### Future directions

Future work may explore if these trends seen in contraceptive provision in Scotland were similar in the other nations of the UK. It is possible the devolved nature of healthcare policy within the UK created disparities in access to contraception during COVID-19 restrictions between the four nations.

Future analysis with more post-COVID-19 data may better analyse the lasting impacts on contraceptive prescribing in Scotland due to COVID-19.

## Data Availability

All code and data used are fully available from Zenodo (doi:10.5281/zenodo.8310085)
The raw dataset used is also publicly available from the Scottish Health and Social Care Open Data repository (opendata.nhs.scot).

https://doi.org/10.5281/zenodo.8310085

## Abbreviations

BNF: British National Formulary
COCP: Combined oral contraceptive pill
Cu-IUD: EC Copper intra-uterine device Emergency contraception
LARC: Long-acting reversible contraception
LNG-IUS: Levonorgestrel intra-uterine system
NHS: National Health Service
POP: Progestogen-only pill
UK: United Kingdom

## Acknowledgements

With thanks to Dr Kasia Banas at the University of Edinburgh, whose excellent data science course inspired this article.

## Reference List

[1] Walker SH. Effect of the COVID-19 pandemic on contraceptive prescribing in general practice: a retrospective analysis of English prescribing data between 2019 and 2020. Contracept Reprod Med 2022;7:3. 10.1186/s40834-022-00169-w.

[2] Balachandren N, Barrett G, Stephenson JM, Yasmin E, Mavrelos D, Davies M, et al. Impact of the SARS-CoV-2 pandemic on access to contraception and pregnancy intentions: a national prospective cohort study of the UK population. BMJ Sex Reprod Health 2022;48:60–5. 10.1136/bmjsrh-2021-201164.

[3] Wood SN, Milkovich R, Thiongo M, Gichangi P, Byrne ME, Devoto B, et al. Disruptions to youth contraceptive use during COVID-19: Mixed-methods results from Nairobi, Kenya. PLOS Glob Public Health 2023;3:e0001005. 10.1371/journal.pgph.0001005.

[4] Roland N, Drouin J, Desplas D, Duranteau L, Cuenot F, Dray-Spira R, et al. Impact of coronavirus disease 2019 on contraception use in France. Therapies 2023. 10.1016/j.therap.2023.01.002.

[5] Cousins S. COVID-19 has “devastating” effect on women and girls. The Lancet 2020;396:301–2. 10.1016/S0140-6736(20)31679-2.

[6] Lewis R, Blake C, McMellon C, Riddell J, Graham C, Mitchell K. Understanding young people’s use and non-use of condoms and contraception. A co-developed, mixed-methods study with 16-24 year olds in Scotland. Final report from CONUNDRUM (CONdom and CONtraception UNDerstandings: Researching Uptake and Motivations) young people’s use and non-use of condoms and contraception 2021. 10.36399/gla.pubs.238377.

[7] Scottish Government. NHS Recovery Plan 2021-2026 2021. https://www.gov.scot/publications/nhs-recovery-plan/documents/ (accessed August 31, 2023).

[8] Wherton J, Greenhalgh T, Shaw SE. Expanding Video Consultation Services at Pace and Scale in Scotland During the COVID-19 Pandemic: National Mixed Methods Case Study. J Med Internet Res 2021;23:e31374. 10.2196/31374.

[9] Grisewood A. Covid-19: Guidance on Planning and Responding to Primary Care GP Practice Capacity Challenges 2020. https://www.sehd.scot.nhs.uk/pca/PCA2020(M)02.pdf (accessed August 31, 2023).

[10] FSRH. Essential Services in Sexual and Reproductive Healthcare 2020. https://web.archive.org/web/20200519182314/ https://www.fsrh.org/documents/fsrh-position-essential-srh-services-during-covid-19-march-2020/ (accessed September 2, 2023).

[11] Geleto A, Taylor J, Beyene T. Interruptions in contraception and unintended pregnancy during the COVID-19 pandemic: A protocol for systematic review and meta-analysis. Womens Health 2023;19:17455057221147382. 10.1177/17455057221147382.

[12] NHS Services Scotland - Information Services Division. Glossary of Terms 2017. http://www.isdscotland.org/Health-Topics/Prescribing-and-Medicines/_docs/Glossary-of-Terms. pdf (accessed September 6, 2023).

[13] R Core Team. R V4.3.0: A Language and Environment for Statistical Computing. 2023.

[14] Joint Formulary Committee. Nonoxinol 2023. https://bnf.nice.org.uk/drugs/nonoxinol/ (accessed August 31, 2023).

[15] Scottish Parliament Information Centre. Timeline of Coronavirus (COVID-19) in Scotland. SPICe Spotlight 2023. https://spice-spotlight.scot/2023/05/10/timeline-of-coronavirus-covid-19-in-scotland/ (accessed September 1, 2023).

[16] Scottish Government. Covid-19 Framework for Decision Making - Scotland’s route map through and out of the crisis 2020. https://www.gov.scot/binaries/content/documents/govscot/publications/strategy-plan/2020/05/coronavirus-covid-19-framework-decision-making-scotlands-route-map-through-out-crisis/documents/covid-19-framework-decision-making-scotlands-route-map-through-out-crisis/covid-19-framework-decision-making-scotlands-route-map-through-out-crisis/govscot%3Adocument/covid-19-framework-decision-making-scotlands-route-map-through-out-crisis.pdf (accessed September 2, 2023).

[17] FSRH Clinical Effectiveness Unit. FSRH Guideline (January 2019, amended November 2020) Combined Hormonal Contraception. BMJ Sex Reprod Health 2019;45:1–93. 10.1136/bmjsrh-2018-CHC.

[18] FSRH. UK MEDICAL ELIGIBILITY CRITERIA FOR CONTRACEPTIVE USE | UKMEC 2016 (AMENDED SEPTEMBER 2019) 2019. https://www.fsrh.org/documents/ukmec-2 (accessed September 3, 2023).

[19] FSRH Clinical Effectiveness Unit. FSRH Guideline (August 2022) Progestogen-only Pills. BMJ Sex Reprod Health 2022;48:1–75. 10.1136/bmjsrh-2022-PoP.

[20] FSRH Clinical Effectiveness Unit. FSRH CEU clinical advice to support provision of effective contraception during the COVID-19 outbreak 2020. https://web.archive.org/web/20200507102018/https://www.fsrh.org/documents/fsrh-ceu-clinical-advice-to-support-provision-of-effective/ (accessed September 4, 2023).

[21] NHS Scotland. National Patient Group Direction (PGD) 326 Supply of Desogestrel Progestogen-Only Contraceptive Pill (POP) Version – 1.0 2021. https://www.communitypharmacy.scot.nhs.uk/media/4913/pgd-326-desogestrel-bridging-contraception-nov-21-final-fillable.pdf (accessed September 6, 2023).

[22] FSRH Clinical Effectiveness Unit. FSRH Guideline (December 2014, amended July 2023) Progestogen-only Injectable Contraception 2014. https://www.fsrh.org/standards-and-guidance/documents/cec-ceu-guidance-injectables-dec-2014/ (accessed September 4, 2023).

[23] FSRH Clinical Effectiveness Unit. FSRH Guideline (February 2021) Progestogen-only Implant. BMJ Sex Reprod Health 2021;47:1–62. 10.1136/bmjsrh-2021-CHC.

[24] Pfizer Limited. SAYANA PRESS 104 mg/0.65 ml suspension for injection SmPC. MedicinesOrgUk 2023. https://www.medicines.org.uk/emc/product/3148/smpc (accessed September 2, 2023).

[25] FSRH Clinical Effectiveness Unit. FSRH Guideline (March 2023) Intrauterine contraception. BMJ Sex Reprod Health 2023;49:1–142. 10.1136/bmjsrh-2023-IUC.

[26] Pfizer Limited. Depo-Provera 150mg/ml Injection Sterile suspension for injection SmPC. MedicinesOrgUk 2023. https://www.medicines.org.uk/emc/product/6721/smpc (accessed September 6, 2023).

[27] MacCrimmon S. Emergency Hormonal Contraception (EHC) Service Service Specification 2015. https://members.cps.scot/media/e5fbbpn3/109813-ehcservicespec_17sep2015.pdf (accessed August 31, 2023).

[28] FSRH Clinical Effectiveness Unit. FSRH Guideline (March 2017, amended July 2023) Emergency Contraception 2017. https://www.fsrh.org/standards-and-guidance/documents/ceu-clinical-guidance-emergency-contraception-march-2017/ (accessed September 6, 2023).

